# Leveraging Pretrained Large Language Model for Prognosis of Type 2 Diabetes Using Longitudinal Medical Records

**DOI:** 10.1101/2025.02.04.24313200

**Authors:** Phong B.H. Nguyen, Andreas Hungele, Reinhard W. Holl, Michael P. Menden

## Abstract

Timely prognosis of type 2 diabetes (T2D) is critical for effective interventions and reducing economic burden. Longitudinal medical records offer potential for extracting clinical insights but face challenges due to the sparse, high-dimensional data, data privacy, domain compatibility and interpretability issues. This study introduced PRIME-LLM, a framework that leverages the prediction power of pretrained large language models for disease prognosis. PRIME-LLM overcomes the challenges by synthetic data generation, missingness modeling, and a learnable embedding layer prepended to a pretrained LLM backbone. We finetuned and evaluated the model performance using a large real world dataset of 449,185 T2D patients. The PRIME-LLM-fine-tuned model outperformed baselines in forecasting HbA1c, LDL, blood pressure, improving MSE up to 12.8%. The model also demonstrated robust long-term prediction over 578.8 days (95% CI: [180,1155]). Integrated gradient analysis identified significant clinical features and visits, revealing potential biomarkers for early intervention. Overall, the results showed the possibility to leverage the prediction power of LLM in T2D prognosis using sparse medical time series, assisting clinical prognosis and biomarker discovery, ultimately advancing precision medicine.

## Introduction

Type 2 diabetes (T2D) is a chronic metabolic disorder that impairs insulin sensitivity and leads to elevated blood glucose level ^1^ and is associated with micro- and macrovascular complications over time, including cardiovascular diseases, neuropathy, nephropathy, and retinopathy, contributing to significant morbidity and mortality worldwide^2^. The rising global prevalence of T2D underscores the urgent need for effective management strategies. Early prognosis of T2D and monitoring the disease trajectory are crucial for enabling timely interventions, reducing risk of complications, improving patient outcomes, and decreasing healthcare costs.

Longitudinal medical records are a vital resource for advancing precision medicine in T2D. These records provide comprehensive clinical presentations over time ^3^, including demographics, medical history, lab results, medications, and lifestyle factors, making them invaluable for early prognosis. Studies like the Framingham Heart Study and DCCT have demonstrated the utility of such data in predicting cardiovascular risk ^4^ and diabetic complications^5^. In the field of diabetes, the DPV registry includes comprehensive biomedical records of hundreds of thousands of diabetes patients across central Europe, representing a uniquely comprehensive resource for T2D studies^6^. These datasets enable the identification of patterns for more accurate and personalized prognostic assessments, but their large volume and complexity pose significant analytical challenges. That being said, research leveraging advanced data analytics to decompose the rich knowledge encompassed in such datasets remains underdeveloped.

Transformer-based deep learning has significantly advanced the time series modeling, especially longitudinal medical records. Several variants of transformers such as Reformer^7^, Informer^8^, Autoformer^9^ and FedFormer^10^ were optimised to increase efficiency and representation learning for general time series forecasting. Med-BERT leverages transformers to encode electronic health records (EHRs), enhancing predictions of disease progression and hospital readmission^11^. Similarly, BEHRT extends the BERT architecture with positional embeddings and masked language model pretraining^12^, enabling detailed patient history representations and improved clinical predictions. While these models have shown substantial performance in clinical representation learning and forecasting, they required relatively large datasets for text embedding and pretraining from scratch, and the transfer learning ability to new tasks and complex datasets of high dimensions and variable sequence lengths is yet to be evaluated.

Large language models (LLMs) have gained tremendous success in natural language processing and computer vision domains, which leads to potential applications in medical time series given the similar autoregressive nature of this kind of data. Particularly, LLMs would offer several advantages over conventional approaches. Their strong generalisability allows transfer across diseases or patient cohorts without retraining from scratch, while their ability to learn from limited samples is valuable in clinical settings where data is scarce. Unlike statistical baselines, LLMs provide conceptual reasoning, enabling recognition of latent physiological patterns that may support more precise prognosis. Their integration of multiple data types (e.g., labs, notes, imaging, signals) opens the door to holistic patient modeling, and their reuse of large-scale pretraining reduces the need for extensive architecture search and hyperparameter tuning that typically burdens medical forecasting models^13^. That being said, the alignment of LLM to time series data exhibits few challenges, as LLM operates on discrete tokens as input, while time series is inherently continuous which makes it not a straightforward task. Moreover, LLMs were pretrained on text, for which their ability to reason and interpret time series data remains an open question.

A few studies have attempted to bridge the gap between the two domains. Meta-Transformer^14^ attempted to unify cross-domain modalities, showing that a single LLM-style transformer could handle tabular, time series, and image-like data. Frozen Pretrained Transformer (FPT)^15^ froze pretrained language transformers and only retrained input/output layers for non-text data, demonstrating that LLM priors could transfer well with minimal tuning. Time-LLM^13^ advanced this by introducing prompt-as-prefix (PaP) reprogramming and data-to-text reformulation, effectively “talking” to LLMs in their native language and achieving strong results in forecasting without full retraining. Similarly, LLMTime^16^ also fine-tuned LLMs on time-series–specific prompts and tasks, enhancing adaptability and performance across diverse datasets. While these methods highlight the promise of reusing powerful LLM priors for time series, their performance in sparse, high dimensional, real world clinical time series is yet to be evaluated. Furthermore, most of them require prompt engineering, text embedding and tokenization which would pose complications in implementation and generalisation. Lastly, interpretation is an important aspect in medical research, but has been less discussed.

This study investigated the application of LLM to medical time series forecasting using a complex longitudinal T2D medical dataset, by proposing a novel approach. We developed PRIME-LLM, a method to directly adapt sparse, structured data as a projected representation suitable for a pretrained large language model (LLM), leveraging minimal preprocessing alongside a learnable embedding layer, eliminating the steps of text processing and tokenization. The model performance was evaluated using synthetic and real world datasets, across several settings and prediction tasks, compared against baselines. Furthermore, the model’s interpretability was also examined, highlighting its potential to support personalized healthcare decision-making. Overall, findings from this study would serve as a proof-of-concept for the utility of LLM in the field of medical forecasting, moving toward personalized medicine for T2D and other diseases.

## Results

This study utilized the German Diabetes Prospective Follow-up Registry (DPV) Initiative ^6^, which contains 649,331 electronic health records from over 400 facilities across four European countries. The analysis targeted 449,185 type 2 diabetes (T2D) patients, selecting 50,183 individuals with at least 10 clinical visits to ensure a balance between sample size and historical data (**Methods**; **Fig. 1a**).

**Figure 1.**
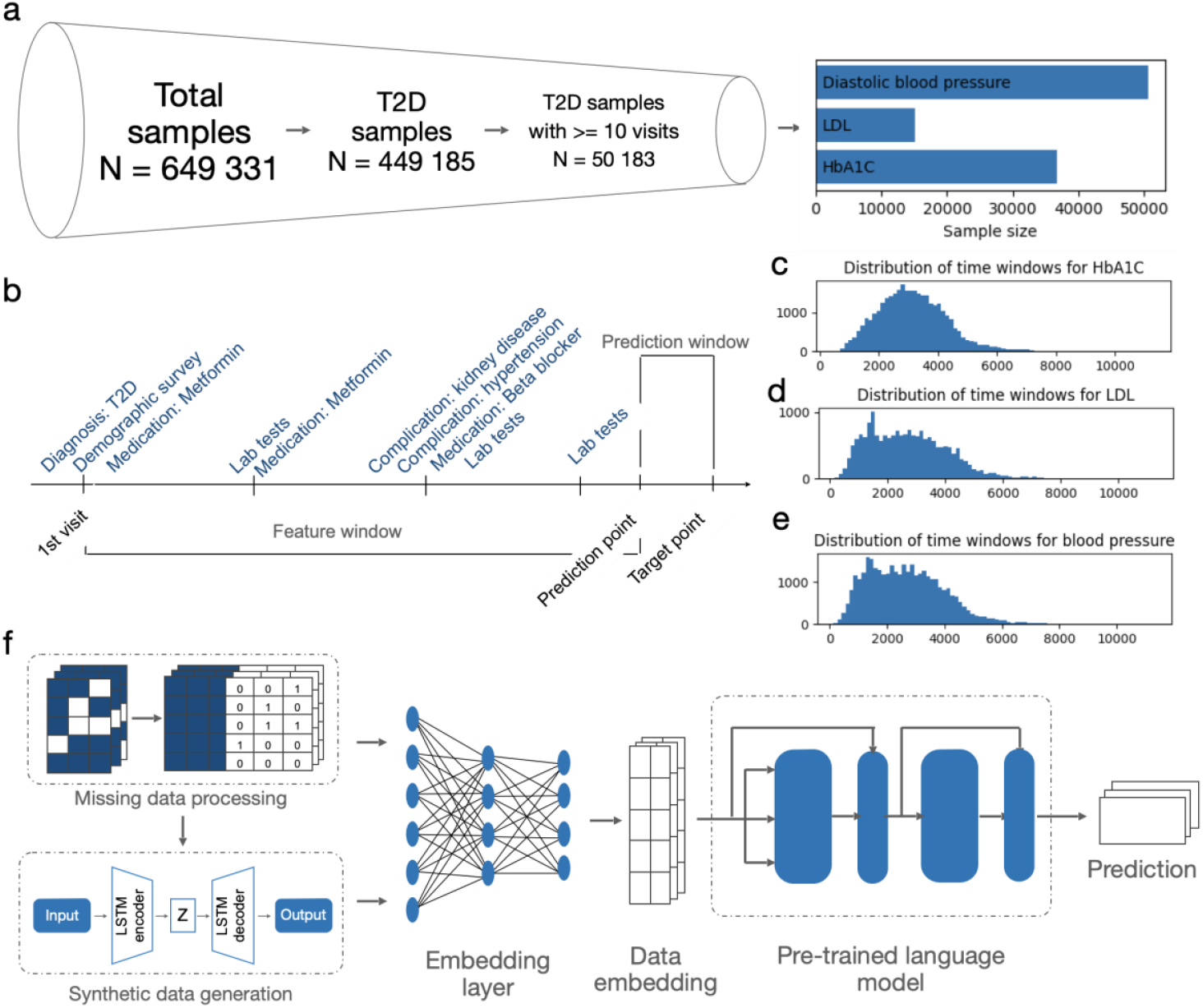
Overview of the dataset and the machine learning framework for adapting sparse medical time series to pretrained large language models. (a) Samples were gone through a filtering process to select relevant samples for each prediction task. The barplots show the numbers of selected, processed samples for HbA1C, LDL and DBP prediction. (b) Example of a patient medical record used in this study, from the first visit to the target point, feature window and prediction window are also defined. (c,d,e) Distributions of the lengths of the time series for HbA1C, LDL and DBP prediction, respectively. (f) Components of the PRIME-LLM framework: We start with processing the sparse medical data to deal with missingness and to generate synthetic data if needed; then implement an embedding layer that outputs a latent representation of the data which is compatible to the hidden space of the pretrained LLM; the embedding is then used as input for the pre-trained language model, which outputs the prediction. During back-propagation, both the embedding and the LLM’s parameters are updated simultaneously.

Each patient’s time series as shown in **Fig. 1b** spanned 10 to 50 visits, capturing up to 321 features (e.g., diagnoses, demographics, measurements, medications, and hospitalizations), with mean missingness of 75%. The mean record period for HbA1c, LDL and DBP prediction samples were 3155 days (95% CI [1410,5145]), 2,691 days (95% CI [840,4955]) and 2665 days [95% CI [890,4905]], respectively (**Fig. 1c,d,e**). After preprocessing, the final datasets comprised 33 746, 18 994 and 31 643 samples for HbA1C, LDL and DBP predictions, respectively (**Fig. 1a**).

### Adaptation of pretrained large language models to sparse multivariate medical time series

The PRIME-LLM framework was developed to process sparse multivariate medical time series data and adapt it for use with pretrained LLMs, harnessing their predictive capabilities in the medical domain. It addresses challenges commonly associated with advanced analytics on medical time series, including missingness, data complexity, domain incompatibility and interpretability, as well as data privacy issues.

The framework consists of multiple features designed to process raw longitudinal data and adapt it to a pretrained LLM (**Methods, Fig. 1f**). The first step is preprocessing, which identifies missing values for each variable using a missing mask. This mask is appended to the original data matrix, and missing values are imputed with the global mean of corresponding variables across time points and samples, ensuring the data remains complete for analysis. By encoding the missingness as additional features, we enable the model to learn the missing patterns and their contribution to the prediction of the target variables. This is especially important in medical prognostics as missing data is often associated with clinical decisions.

As data privacy issues are common in biomedical research, PRIME-LLM also offers a solution to generate synthetic data to tackle this problem. Specifically, we implement an in-house LSTM-VAE pipeline which can learn the underlying representation of the original data and reconstruct it in completely synthetic data (**Methods, Fig. 1e**). By using this feature, we could increase the training sample size and train machine learning models without violating data privacy issues.

Once preprocessing is complete, the data is passed through an embedding layer (**Methods, Fig. 1e**). This linear layer alters the dimensionality of the data while preserving its essential features, ensuring the data is compatible with the embedding layer of the pretrained LLM that will be used for prediction. Overall, the missingness processing and embedding layer enable the direct plugin of raw medical time series to LLMs without having to transform into text and engineer prompts for providing context. We hypothesized that the pretraining of LLMs provided them the ability to learn complex mathematical relationships of new data and domains without explicit context provision.

In the final step, the embedded data is input into a pretrained LLM, which has been trained on vast amounts of textual data, before a final linear layer for the output. The LLM is adapted to predict health outcomes based on the provided structured medical data (**Methods, Fig. 1e**). In this study, the framework was used to predict HbA1c, LDL and DBP levels for either the next visit or a future visit, demonstrating its ability to predict complex health outcomes from longitudinal medical records.

### PRIME-LLM exhibited strong performance even with limited sample sizes

To access PRIME-LLM prediction performance and transfer learning ability, we utilized a synthetic dataset generated based on the original DPV data structure and distributions, using our in-house LSTM-VAE method, incorporated into the PRIME-LLM (**Methods**). Several training sizes were used to evaluate the models, as described in **Methods**. The target variable to predict in this benchmark is HbA1C. Since the model aimed to support therapeutic strategy, we excluded HbA1C and any variables that were strongly associated with it in the modeling process.

We used various baselines to compare against our method. Specifically, we involved non-deep learning algorithms such as elastic net and XGBoost to evaluate the ability of deep learning to learn complex horizontal and longitudinal relationships. Autoregressive deep learning architectures such as LSTM, vanilla Transformer, Autoformer and FEDforme were also implemented in the benchmark to evaluate the advantage of cross-modality transfer learning by LLMs. Finally, we tested several LLM backbones for PRIME-LLM, including BERT base, GPT-2, TinyLlama and Phi-1.5, which are different from each other in the number of parameters, transformer architectures and pretraining process.

Overall, PRIME-LLM showed strong performance, ranked either first or second in terms of mean square error (MSE) across most training sizes, especially for very low numbers of samples (**Table 1 & Supp.Fig.S1**). Particularly, PRIME-LLM which used TinyLlama and Phi-1.5 as backbone achieved superior performance, ranked first and second, respectively for training sizes of 200 samples or less. At train size of 50, PRIME-LLM-TinyLlama and -Phi-1.5 decreased MSE by 40.6% and 39.7%, respectively, compared to Autoformer, the best performer at that size. At train size of 200, the two models decreased MSE by 13.7% and 13.2%, respectively, compared to elastic net, the best performer at that size. Furthermore, smaller LLMs such as BERT and GPT-2 still outperformed other deep and non-deep learning methods in training sizes of 50 and 100, although their performance was not comparable to TinyLlama and Phi-1.5. This highlighted the importance of LLM choices when implementing PRIME-LLM, as larger LLMs tend to perform better. Overall, this demonstrated the applicability of PRIME-LLM to this data context, and the transfer learning ability of LLM, as only a few samples were required to fine tune the model in a new dataset and context, in order to achieve strong performance.

**Table 1.** Prediction performance of different machine learning algorithms across different training sizes of the synthetic data. For each algorithm, random subsets of 25 000 samples were used as training sets, the rest were used as test sets for performance evaluation. The sampling process was repeated five times per algorithm per training size. Recorded values here are in the format of “mean [95% CI]” of the mean squared error. The best performing model for each training size is in red, the second best is in blue.

|  | Train size (out of 25 000 samples) |  |  |  |  |  |  |  |  |  |  |
| --- | --- | --- | --- | --- | --- | --- | --- | --- | --- | --- | --- |
| Model | 50 | 100 | 200 | 500 | 1000 | 2000 | 5000 | 7500 | 10000 | 15000 | 20000 |
| Elastic net | 0.1752 [0.1454,0.2051] | 0.1264 [0.0947,0.1582] | 0.0840 [0.0553,0.1128] | 0.0463 [0.0432,0.0495] | 0.0376 [0.0346,0.0406] | 0.0295 [0.0269,0.0322] | 0.0231 [0.0209,0.0253] | 0.0213 [0.0195,0.0232] | 0.0207 [0.0193,0.0221] | 0.0199 [0.0182,0.0216] | 0.0182 [0.0157,0.0208] |
| XGBoost | 0.1596 [0.1347,0.1846] | 0.1207 [0.1054,0.1360] | 0.0848 [0.0738,0.0957] | 0.0426 [0.0381,0.0472] | 0.0302 [0.0281,0.0322] | 0.0213 [0.0195,0.0231] | 0.0142 [0.0128,0.0156] | 0.0113 [0.0105,0.0121] | 0.0103 [0.0095,0.0111] | 0.0078 [0.0069,0.0087] | 0.0073 [0.0051,0.0094] |
| LSTM | 0.2638 [0.1669,0.3608] | 0.2066 [0.1969,0.2163] | 0.1921 [0.1880,0.1961] | 0.1731 [0.1674,0.1788] | 0.1650 [0.1609,0.1692] | 0.1567 [0.1533,0.1602] | 0.1534 [0.1459,0.1609] | 0.1445 [0.1398,0.1492] | 0.1414 [0.1383,0.1445] | 0.1320 [0.1258,0.1382] | 0.1350 [0.1263,0.1436] |
| Transformer | 0.1372 [0.1276,0.1468] | 0.1362 [0.1217,0.1506] | 0.1282 [0.1171,0.1392] | 0.1127 [0.1081,0.1173] | 0.0944 [0.0902,0.0987] | 0.0731 [0.0690,0.0773] | 0.0471 [0.0416,0.0525] | 0.0369 [0.0342,0.0396] | 0.0297 [0.0283,0.0312] | 0.0219 [0.0200,0.0237] | 0.0205 [0.0160,0.0249] |
| Autoformer | 0.1304 [0.1246-0.1343] | 0.1291 [0.1204-0.1372] | 0.116 [0.1105-0.1237] | 0.1048 [0.1004-0.1094] | 0.0998 [0.097-0.1048] | 0.0842 [0.0811-0.0888] | 0.069 [0.0665-0.0743] | 0.0639 [0.0594-0.0739] | 0.0626 [0.0611-0.0643] | 0.0652 [0.0600-0.0748] | 0.0732 [0.0543-0.0918] |
| FEDformer | 0.1260 [0.1186,0.1334] | 0.1302 [0.1195,0.1408] | 0.1198 [0.1088,0.1308] | 0.1122 [0.1088,0.1156] | 0.1053 [0.0958,0.1148] | 0.1140 [0.0789,0.1491] | 0.1161 [0.0693, 0.1628] | NaN | NaN | NaN | NaN |
| PRIME-LLM-BERT | 0.1298 [0.1253-0.1353] | 0.129 [0.1232-0.1354] | 0.118 [0.1143-0.1217] | 0.1022 [0.0975-0.1076] | 0.083 [0.08-0.0856] | 0.0623 [0.0607-0.0634] | 0.0388 [0.0374-0.0406] | 0.0306 [0.0298-0.0313] | 0.027 [0.0252-0.0289] | 0.0215 [0.0196-0.0235] | 0.0194 [0.0182-0.0203] |
| PRIME-LLM-GPT2 | 0.1006 [0.0909,0.1104] | 0.0959 [0.0904,0.1014] | 0.0896 [0.0856,0.0937] | 0.0727 [0.0696,0.0757] | 0.0610 [0.0582,0.0639] | 0.0466 [0.0447,0.0484] | 0.0292 [0.0275,0.0309] | 0.0235 [0.0221,0.0249] | 0.0212 [0.0203,0.0221] | 0.0178 [0.0162,0.0193] | 0.0165 [0.0151,0.0180] |
| PRIME-LLM-TinyLlama | 0.0775 [0.0746-0.0814] | 0.0768 [0.0734-0.0819] | 0.0725 [0.0696-0.0758] | 0.0607 [0.0576-0.0631] | 0.0518 [0.0508-0.0531] | 0.0388 [0.0376-0.04] | 0.0252 [0.0245-0.0262] | 0.0204 [0.0195-0.0214] | 0.0181 [0.0169-0.0199] | 0.0147 [0.0129-0.0167] | 0.0144 [0.0122-0.0177] |
| PRIME-LLM-Phi1.5 | 0.0786 [0.0770,0.0803] | 0.0778 [0.0752,0.0804] | 0.0729 [0.0707,0.0751] | 0.0621 [0.0584,0.0657] | 0.0498 [0.0483,0.0514] | 0.0368 [0.0353,0.0384] | 0.0239 [0.0221,0.0256] | 0.0118 [0.0117,0.0120] | 0.0094 [0.0037,0.0152] | 0.0083 [0.0072,0.0095] | 0.0068 [0.0056,0.0081] |

There was a variation in performance among other methods in the benchmark. Vanilla Transformer, Autoformer and FEDformer, showed comparable performances, as they outperformed LSTM, XGBoost and elastic net at train sizes of 50 and 100, suggesting the efficiency of these transformer-based architectures in learning the underlying time series representation, which simpler algorithms such as XGboost and elastic net were not inherently designed for. However, XGBoost and elastic net overtook at higher training sizes, suggesting the effectiveness of the traditional machine learning models when they have enough data to account for the high dimensions of the dataset. On the other hand, LSTM exhibited poor performance when compared to either simpler architectures or more efficient learning algorithms.

### PRIME-LLM successfully forecasted metabolic indicators in type 2 diabetes patients

Given the promising performance on the synthetic data, we moved on to evaluate PRIME-LLM on a real world T2D dataset. In particular, we benchmarked PRIME-LLM-Phi-1.5, which consistently performed well in the previous analysis, with XGBoost and elastic net, the two good performing traditional machine learning algorithms, and a vanilla transformer. Implementing these methods, we attempted to predict HbA1C, LDL and DBP in the next doctor visits from the observed time series, as these are common and important metabolic indicators for several conditions, including T2D and its complications. The data sampling and cross validation design were similar to that of the synthetic data, as detailed in **Methods**.

The PRIME-LLM-Phi-1.5 models outperformed other models in all three prediction tasks at train size of 50 samples, while consistently ranked high large training sizes **(Table 2 & Supp.Fig.S2**), which was in accordance with what was observed in the synthetic data. Particularly, at train size of 50, compared to Transformer-the second best performer, PRIME-LLM-Phi-1.5 decreased the MSE by 12.8% for HbA1C, 3.3% for LDL and 1.1% in DBP. In addition, the model also maintained good performance at larger train sizes. For HbA1C prediction, it decreased the MSE by 1.9% compared to the second best performer XGBoost at 20,000 samples. For LDL at 12000 sample train sets, it decreased MSE by 1.6% compared to the transformer model. For DBP prediction, PRIME-LLM-Phi-1.5 model ranked second at 20000 samples, only outperformed by the XGBoost model. Overall, although real world data represents more challenges compared to synthetic data, our approach still executed all the prediction tasks, reflected in the excellent performance compared to the baselines.

**Table 2.** Prediction performance of different machine learning algorithms across different training sizes and prediction tasks on the real world dataset. For each algorithm, random subsets of 25 000 samples were used as training sets, the rest were used as test sets for performance evaluation. The sampling process was repeated five times per algorithm per training size. Recorded values here are in the format of “mean [95% CI]” of the mean squared error. The best performing model for each training size is in red, the second best is in blue.

|  | Train size (out of 25 000 samples) |  |  |  |  |  |
| --- | --- | --- | --- | --- | --- | --- |
|  | HbA1C |  | LDL |  | Diastolic blood pressure |  |
| Models | 50 | 20,000 | 50 | 12,000 | 50 | 20,000 |
| Elastic net | 1.0027<br>[0.8733,1.1321] | 0.6563<br>[0.6320,0.6807] | 3.9854<br>[3.7413,4.2296] | 3.6032<br>[3.4431,3.7633] | 1.9989<br>[1.7487,2.2492] | 1.6338<br>[1.5882,1.6795] |
| XGBoost | 0.9075<br>[0.7843,1.0307] | **0.5861**<br>**[0.5761,0.5960]** | 4.5410<br>[4.2186,4.8634] | 3.4883<br>[3.3097,3.6668] | 2.1120<br>[1.6976,2.5264] | **1.5537**<br>**[1.5107,1.5967]** |
| Transformer | **0.7920**<br>**[0.6345,0.9496]** | 0.6556<br>[0.6270,0.6842] | **3.6948**<br>**[3.6161,3.7735]** | **3.1804**<br>**[3.0949,3.2659]** | **1.6901**<br>**[1.6460, 1.7500]** | 1.5888<br>[1.548, 1.642] |
| PRIME-LLM-Phi1.5 | **0.6908**<br>**[0.6494,0.7322]** | **0.5751**<br>**[0.5526,0.5976]** | **3.5743**<br>**[3.4644,3.6841]** | **3.1288**<br>**[3.0316,3.2259]** | **1.6709** [1.629,<br>1.745] | **1.5798**<br>**[1.5311,1.6284]** |

### LLM-based models excel in long-term medical time series forecasting

Long term time series forecasting has a practical application in the medical field, which motivated us to evaluate the clinical utility of our models in such circumstances. To this end, we compared the PRIME-LLM-Phi-1.5 performance when predicting the next visit, against the cases when the target points were further away in the future. Specifically, the comparison focused on predicting HbA1c levels at varying prediction window lengths by moving the prediction points backward, ranging from a mean of 97.1 days (95% CI: [15, 195]) to 1313.9 days (95% CI: [420, 2895]) (**Fig. 2a**). To have a more comprehensive result, we also involved the XGBoost model in the benchmark, evaluated at 20000 samples in training sets.

**Figure 2.**
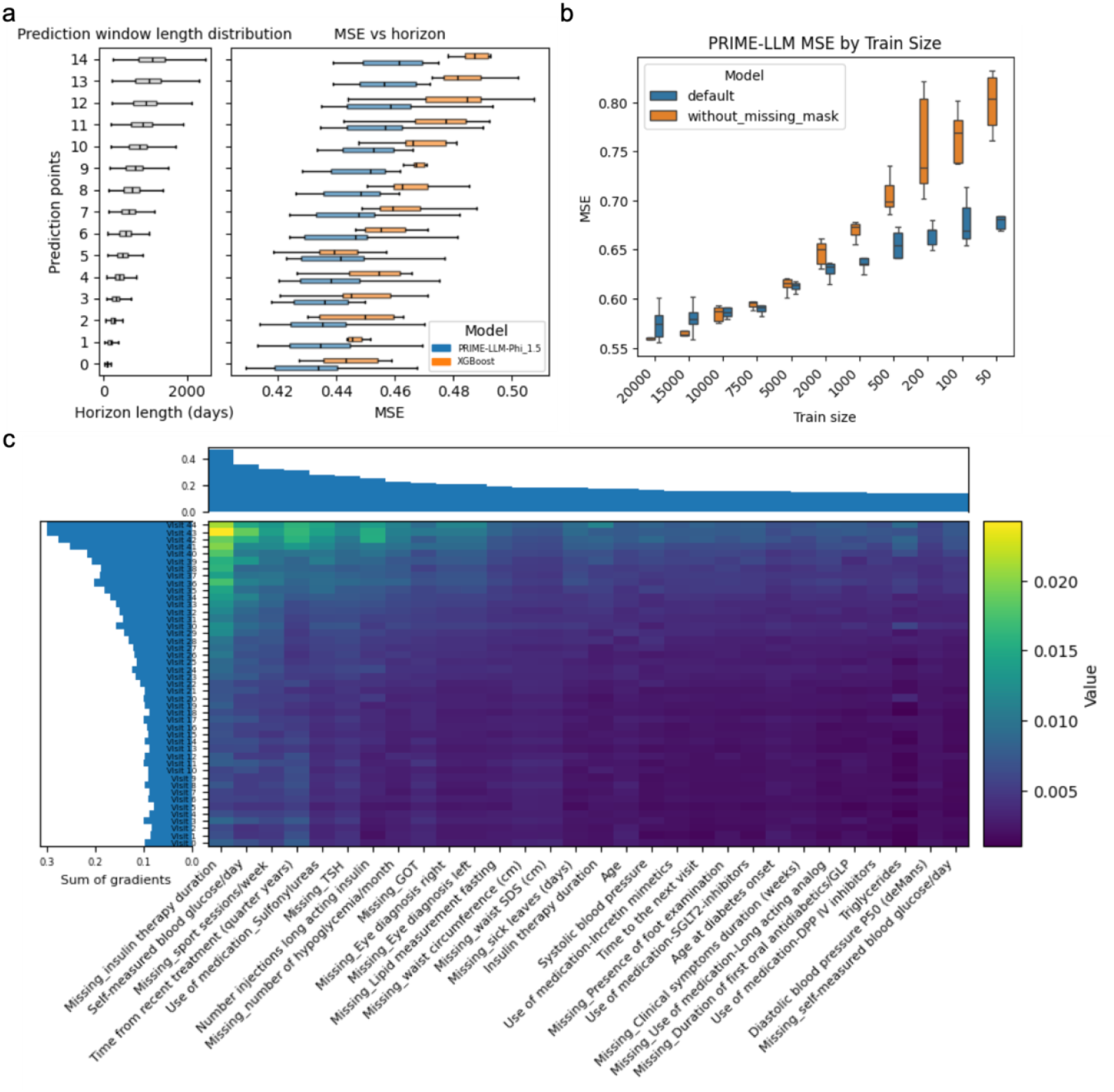
Prediction performance across different horizons, utility of the missing mask and interpretability. a) Horizon distribution at different prediction points (left) and the prediction performance of PRIME-LLM-Phi-1.5 and XGBoost at different horizon windows (right). Results were recorded for 10 resamples, each consisted of 20 000 samples in training and 5000 for testing. b) Ablation study comparing the prediction performance across different training sizes of the PRIME-LLM-Phi-1.5 default and PRIME-LLM-Phi-1.5 without the appended missing mask. Results were recorded for 5 resamples. c) The feature importance scores of HbA1C prediction, represented by the integrated gradients of the top 30 most important features (columns) across all time points (rows). The values represent the mean absolute gradients of the features across a subset of 1000 samples.

The PRIME-LLM-Phi-1.5 was comparable to XGBoost in prediction performance at shorter horizons, but the performance differences became evident in longer term predictions (**Fig. 2a**). In particular, for our model, the MSE at prediction point 7 where the horizon was 578.8 (95% CI: 180, 1155]) was not significantly different from that at prediction point 0 (97.1 days (95% CI: [15, 195])), (Wilcoxon Rank Sum test, p=0.273), only at prediction point 14 where the horizon was 1313.9 days (95% CI: [420, 2895]) did the different become significant (p=0.031). In contrast, XGBoost decreased the performance significantly at both prediction point 7 and 14 compared to point 0 (p=0.021 and p=4.4e-04, respectively). Overall, these results suggested the robustness of our model in long term prediction, allowing more room for clinical intervention.

### The feature missing mask is essential for the robust performance of PRIME-LLM

To evaluate the importance of using the missing mask as an added feature of PRIME-LLM to learn the missing pattern of our data, we conducted an ablation study in which two versions of PRIME-LLM were compared against each other: default setting with the missing mask and a customised framework removing the missing mask. When benchmarking across different training sizes to predict HbA1C, the default PRIME-LLM-Phi-1.5 showed superior performance against the ablated model, especially at lower training sizes. Specifically, at larger sample sizes such as 20 000 and 15 000, the two showed comparable prediction MSE, but starting from 2000 down to 50 samples, the PRIME-LLM with missing mask significantly outperformed the one without (p < 0.05, Wilcoxon Rank Sum test) (**Fig. 2b**). This highlighted the essentiality of the missing mask in assisting the LLM in learning the data features, especially when the data was scarce and the feature space was large.

### PRIME-LLM potentially assists clinical decision making by enhancing interpretability for prognosis of T2D

To better understand the clinical relevance of the PRIME-LLM models in studying T2D trajectory, we performed an interpretability analysis. Specifically, we focused on identifying the features and time points most influential for predictions of the metabolic indicators. Using integrated gradients, a robust method for evaluating feature importance (**Methods**), we analyzed the gradients of a subset of samples across 45 doctor visits.

The feature analysis for HbA1C prediction provided valuable insights into the temporal aspects influencing the model’s decision-making. Most of the feature importance gradients were concentrated on the more recent visits before the target point, indicating that the information from these visits is crucial for accurate prediction (**Fig. 2c**). This suggests that recent medical history plays a significant role in predicting future HbA1c levels, although distant history should also be taken into account.

In terms of specific features, the analysis revealed several important clinical information that was influential for the final HbA1C value, as indicated by the highest absolute gradients across samples and time points (**Fig. 2c**). Common factors such as age, blood pressure and triglycerides appeared on the list, reinforcing the importance of these risk factors and the sensitivity of our model. Among the top influencers, treatment information represented a major group, as insulin therapy duration, type and frequency, and the use of certain medications such as sulfonylureas, incretin mimetics, SGLT2 inhibitors and DPP4 inhibitors play important roles in regulating HbA1C level.

The lack of recording of certain information such as number of sport sessions per week, TSH level, number of hypoglycemia episodes, eye diagnosis and many physiological measurements seemed to be associated with the final HbA1C level. This suggested that the missing patterns could be used as indirect indicators of metabolic conditions, and called for more research to investigate the association and identify the causal factors.

## Discussion

In this work, we introduced PRIME-LLM, a novel approach to adapt sparse numerical data from medical time series to a pretrained large language model (LLM), exemplifying by making predictions on the DPV dataset. Instead of using traditional text prompt engineering, we modeled missing patterns and added a learnable embedding layer between the input and pretrained LLM layers. This approach allowed direct input of numerical data into the transformer architecture, effectively and efficiently leveraging the LLM’s predictive power. Our results demonstrate strong performance in both prediction and clinical utility.

The PRIME-LLM demonstrated strong predictive performance for synthetic and real world data in tasks predicting common metabolic indicators including HbA1C, LDL and DBP, outperforming baseline models across various training sizes, with robust results even at smaller sample sizes. This advantage is likely due to the LLM’s pretraining on large datasets, reducing the need for extensive fine-tuning and allowing transfer learning to new data modality and context. Although the performance improved significantly with larger sample sizes, PRIME-LLM shone at smaller sample sizes, exhibiting the ability to efficiently learn the data context and make predictions even with less than 200 samples, given the vast dimensionality of the real world dataset. Overall, these results suggested the specific advantage of PRIME-LLM for sparse, multivariate medical time series prediction, in the area where conventional deep and non-deep learning methods are struggling.

One strength of our method is the ability to make long term predictions. The result showed that PRIME-LLM robustly predicted HbA1C levels up to 20 months ahead, showing no significant decrease in prediction performance. This highlighted the robustness of our method in capturing distant clinical information to make long term predictions. Furthermore, this also showed the importance of historical data in prediction, even though the most recent visits were more important. Overall, the approach shows strong clinical utility for type 2 diabetes prognosis.

An important aspect of our model is its ability to provide mechanistic insights into the individual patient’s pathological trajectory. To enhance accessibility for patients and healthcare practitioners, interpretability is essential. In this study, we used integrated gradients to interpret the model’s predictions, focusing on important visits and influential features. Our analysis suggests that while historical data is important, the most recent visits hold the most relevance for prognosis. Furthermore, our direct end-to-end architecture provided a straightforward way to identify both global features and signatures associated with individual patients. This enabled detailed investigation into model decision making. Overall, this highlights the model’s clinical utility in guiding patient care decisions.

Another key advantage of our study is the use of a large longitudinal T2D patient dataset from the DPV registry. While longitudinal health data has shown great potential in medical applications for T2D by harbouring a vast space of patient clinical features, leveraging the prediction power of LLMs on such datasets is unprecedented. Our findings are a proof of concept for the utility of LLM for the prognosis of T2D, overcoming data privacy issues and analytical challenges.

PRIME-LLM models demonstrate strong clinical performance but face limitations. These models rely on pretrained LLMs, which come with a wide range of pretraining techniques, numbers of parameters and underlying architectures. Larger causal LLM tended to perform better, but required substantial computational resources. Thus, careful data preprocessing, fine-tuning, optimisation and resource considerations are still needed to adapt the method to specific context. Furthermore, the performance was evaluated solely on clinical data, which might limit the generalisability to other data contexts. Future studies that integrate more data modalities such as omics data, images and clinical notes would reveal more insights into its performance.

In conclusion, this study introduced a novel approach leveraging pretrained LLMs for the prognosis of T2D patients using sparse, complex medical time series. The PRIME-LLM method demonstrated superior performance in forecasting future HbA1c, LDL and DBP levels, identifying critical past visits, and highlighting influential features as potential biomarkers for these metabolic indicators. These results underscore the clinical utility of the approach in enhancing diagnosis, prognosis, and prevention strategies, thereby advancing the application of precision medicine in healthcare.

## Methods

### Study dataset, data processing and experimental design

The study was conducted under a data sharing agreement with the DPV Initiative, a project initiated by the German Diabetes Center (DZD) and executed and maintained by Ulm University, Germany, from which we obtained the data. The DPV Initiative has been collecting patient medical records since the 1990s from over 500 treatment facilities across Germany, Austria, Switzerland and Luxembourg. At the time of acquisition, the data consisted of 649,331 patients, of which 449,185 were of T2D. The number of doctor visits in T2D patients ranged from 1 to 241.

To avoid large sequence gaps and ensure sufficient data for model training, patient records were segmented into sequences of length *n* (10–50). Each timeline consisted of three parts: (1) the **feature window** (t_1_–t_n−1_), covering all visits up to the prediction point and including up to 321 features per visit, though not all features were recorded at every visit; (2) the **prediction point** (t_n−1_), representing the last observed visit used for forecasting; and (3) the **prediction window** (t_n−1_–t_n_), which defines the forecasting horizon, with the target point (t_n_) containing the outcome to be predicted.

The final numbers of samples were 33 746 for HbA1c, 18 994 for LDL and 31 643 for DBP. The dataset initially comprised 321 variables, encompassing demographics, lifestyle factors, physiological measurements, diagnoses, treatments, medications, and administrative records. These variables underwent extensive preprocessing to ensure data quality and compatibility with the model. Uniform variables, those entirely missing, and variables with less than 5% data availability were excluded. Inconsistent encodings were corrected, and highly correlated or redundant variables were removed. Categorical variables were transformed using one-hot encoding.

### Synthetic data generation

To augment the limited amount of real patient data and facilitate the benchmarking, we employed an LSTM with Variational Embedding and Attention (LSTM-VAE) model to generate a synthetic dataset. Specifically, the LSTM backbone was trained to capture the temporal dynamics of our multivariate medical time series, while the variational embedding introduced stochasticity to represent patient-level variability and reduce overfitting. After training, the model was used in generative mode: sampling from the latent embedding space and decoding through the LSTM pipeline to produce synthetic sequences that preserve the statistical properties and temporal patterns of the original dataset while introducing realistic diversity. This synthetic dataset was then used for model training and evaluation.

### The PRIME-LLM framework

The PRIME-LLM consists of three main components:

1. **Missingness processing:** To address missing data, numerical variables were imputed using the global mean of the respective variable, while missing values in categorical variables were encoded as “missing” prior to one-hot encoding. Additionally, a missing mask was generated, capturing the missingness pattern by encoding ‘1’ for missing and ‘0’ for present data points. This mask, designed to retain potentially informative missingness patterns, was appended to the processed data tensor. In addition, to account for the variable sequence lengths, we padded any time series that has n < 50 with 0 and provided a padding tensor which served as the attention mask that guided the LLM to focus on the observed time steps.
2. **Embedding:** To enable time series data integration with a pretrained LLM without requiring text prompt engineering, we implemented a learnable linear embedding layer that transformed raw numerical inputs into latent representation that both captured the relevant patterns in the data and acted as an adapter that provided data context to the LLM. The embedding component was designed as a multilayer perceptron (MLP) whose output was compatible with the adapted LLM.
3. **Pretrained LLM:** The last component was a pretrained LLM that took the output from the embedding component, together with the attention mask, as its input directly into its internal embedding layer, making an end-to-end framework. The LLMs were initiated with the pretrained parameters provided through platforms such as HuggingFace^17^. Most of the internal architecture was kept intact. We only appended a linear output layer at the end after the hidden state of the last token, which outputted a single value, to be compatible with our forecasting tasks. In addition, to assist the fine-tuning of relatively larger LLMs such as TinyLlama and Phi-1.5 while having limited resources, we implemented Low-Rank Adaptation (LoRA)^18^, an efficient way to fine-tune LLM without updating all of its parameters.

We optimized the framework’s several hyperparameters, including specific parameters from PRIME-LLM, the pretrained LLM and LoRA, using Bayesian Optimisation^19^. Details of the optimisation process is in section **Optimisation and training**, and the list of all optimised parameters could be found in **Supp. Data S1**.

### Implementation of elastic net

In order to implement elastic net with the processed time series data, we first flattened the data into a table of shape [s, n*p] where s, n and p were number of samples, sequence length and number of features, respectively. Next we removed the low variance feature with a variance threshold of 1e-05, and selected the top 2000 features based on the mutual information with the target variable. The feature selection and model training were performed using the scikit-learn library^20^. Details of the optimisation process is in section **Optimisation and training**, and the list of all optimised parameters could be found in **Supp. Data S1**.

### Implementation of XGBoost

We implemented a similar data flattening procedure and feature selection procedure as in elastic net. The XGBoost models were trained using the XGBRegressor function in the Python xgboost library^21^. The training and testing scheme followed the same protocol as the LLM- and LSTM-based methods. Details of the optimisation process is in section **Optimisation and training**, and the list of all optimised parameters could be found in **Supp. Data S1**.

### Implementation of long short term memory (LSTM)

The LSTM model took the processed data and its dimension as input and input dimension. The model consists of several units corresponding to the time steps, each consisting of an optimizable number of layers and hidden dimensions, and a linear output layer at the end to provide final prediction. The model parameters were initiated from scratch using Pytorch library. Details of the optimisation process is in section **Optimisation and training**, and the list of all optimised parameters could be found in **Supp. Data S1**.

### Implementation of Transformer

The Transformer for our time series was a simplified encoder-decoder architecture designed for regression tasks. The input, timestamps of the sequence and the attention mask were plugged directly into the model. Each timestep is projected into a hidden dimension, with timestamp encodings added to retain order and time information. The model uses a standard Transformer encoder with encoder blocks consisting of attention heads and a number of layers, and feedforward networks. The decoder is simplified to a linear projection from the last hidden state, predicting the next timestep). The model parameters were initiated from scratch using Pytorch library. Details of the optimisation process is in section **Optimisation and training**, and the list of all optimised parameters could be found in **Supp. Data S1**.

### Implementation of Autoformer

Autoformer^9^ was designed specifically to tackle long-term time series forecasting problems. Its core innovation is the Auto-Correlation mechanism, which replaces the standard self-attention. Instead of directly attending to all positions (which is quadratic in cost), Autoformer explicitly learns periodic dependencies in the data by selecting time-delay correlations and aggregating across them. On top of this, Autoformer introduces a series decomposition block that separates the input into trend and seasonal components. Each part is processed independently, and then recombined, which improves stability and interpretability compared to treating the series as a raw signal. We obtained the official model architecture as described in the paper. Since there was no option to input an attention mask, we padded the shorter sequences with the last observed values, to account for the variable sequence lengths.

### Implementation of FEDformer

FEDformer^22^ builds on Autoformer but goes one step further by introducing frequency-domain attention. Instead of operating purely in the time domain, FEDformer projects the series into either Fourier or Wavelet space, and then applies attention only to a small number of important frequency modes. This is called Frequency Enhanced Attention (FEA). By focusing on low-frequency components (long-term structures) and reducing redundant high-frequency interactions, FEDformer achieves both lower computational cost and better forecasting accuracy. We obtained the official model architecture as described in the paper. Since there was no option to input an attention mask, we padded the shorter sequences with the last observed values, to account for the variable sequence lengths.

### Optimisation and training

We implemented Bayesian Optimization (BO) to optimize the hyperparameters, implemented using the Optuna library^23^. BO is a global optimization strategy that uses a probabilistic surrogate model (like a Gaussian process or Tree Parzen Estimator) and an acquisition function to efficiently balance exploration and exploitation when searching for the best hyperparameters. For each model training round, before the cross-validation, we reserved a subset of data for BO, usually 1000 samples. The optimization consisted of 50 trials, each consisted of 800 samples for training and 200 samples for validating. Each trial would use the validating result from the previous ones to search for the next set of optimal hyperparameters.

The maximum number of epochs for training was 200, with early stopping after 30 epochs of not improving in the validation, to prevent over-fitting. After 50 trials, the best set of parameters was obtained and used for the cross-validation to evaluate the model performance. List of all optimized parameters is in **Supp. Data S1**.

The cross-validation process consisted of five or ten resamples. In each resample, the dataset was split into train, validate (for deep learning models) and test sets, at different ratios depending on the analysis. The models were initialized with the optimized hyperparameters. Training was also done at maximum 200 epochs with early stopping as described above. Adam Optimizer with optimized learning rate was used, with mean square error as the metrics. To prevent over-fitting, we also implemented dropout and weight decay which were also optimized in the previous step.

### Benchmarking with different training sizes

We used a set of 25 000 samples of either synthetic or real data for this analysis (15 000 in case of LDL prediction, due to small sample size). Within the selected samples, prediction performance was assessed across various training set sizes, including 50, 100, 200, 500, 1000, 2000, 5000, 7500, 10 000, 15 000, and 20 000 samples. The remaining samples were reserved for testing.

For deep learning models, 20% of samples from the training set were allocated for validation. The maximum number of epochs for training was 200, with early stopping after 30 epochs of not improving in the validation, to prevent over-fitting. Models were evaluated on 5 or 10 resampled datasets for each train-test ratio.

### Prediction Window Creation and Evaluation

Prediction windows were generated by shifting the prediction point backward along the patient’s timeline. Specifically, the prediction point was moved from the visit immediately preceding the target point up to 15 visits prior, resulting in corresponding prediction windows or horizons. For each window, the same training and evaluation procedure as described for benchmarking training set sizes was applied.

### Integrated gradients and analysis of feature importance

We used integrated gradients to interpret the model predictions. This feature attribution method quantifies the contribution of each input feature by calculating the path integral of gradients from a baseline input (typically zero) to the actual input. It computes the gradients of the model’s output with respect to the input at various points along this path and averages them. Integrated gradients offer a more stable and accurate attribution of feature importance compared to simpler gradient methods. Further details can be found in the original publication^24^.

To interpret the model predictions, we computed integrated gradients for a subset of 1000 samples with 45 visits using the trained model. Both total gradients and average absolute gradients across all samples were calculated. The average absolute gradients were then used to identify influential visits, and the most important individual features were determined by summing the gradients across time points.

### Statistical and computational analysis of results

The analysis framework was developed using PyTorch, with data processing, model training, and evaluation conducted using various Python libraries. Hypothesis testing for two continuous variables was performed using the Wilcoxon Rank Sum test, with p-values adjusted for FDR. For two categorical variables, Fisher’s Exact test was used. Data visualization was carried out using ggplot2 in R and matplotlib in Python.

## Code availability

The code for the analysis and computational framework is located here: https://github.com/phngbh/PRIME_LLM.

## Data availability

The data is not publicly accessible due to the patient’s confidentiality. Researchers who would like to reproduce the results could contact the data access committee at the DPV Initiative in Ulm University. Detailed information can be found here: https://buster.zibmt.uni-ulm.de/.

