## Supplementary figures and images for "Leveraging Pretrained Large Language Model for Prognosis of Type 2 Diabetes Using Longitudinal Medical Records"

### Supplementary Figures

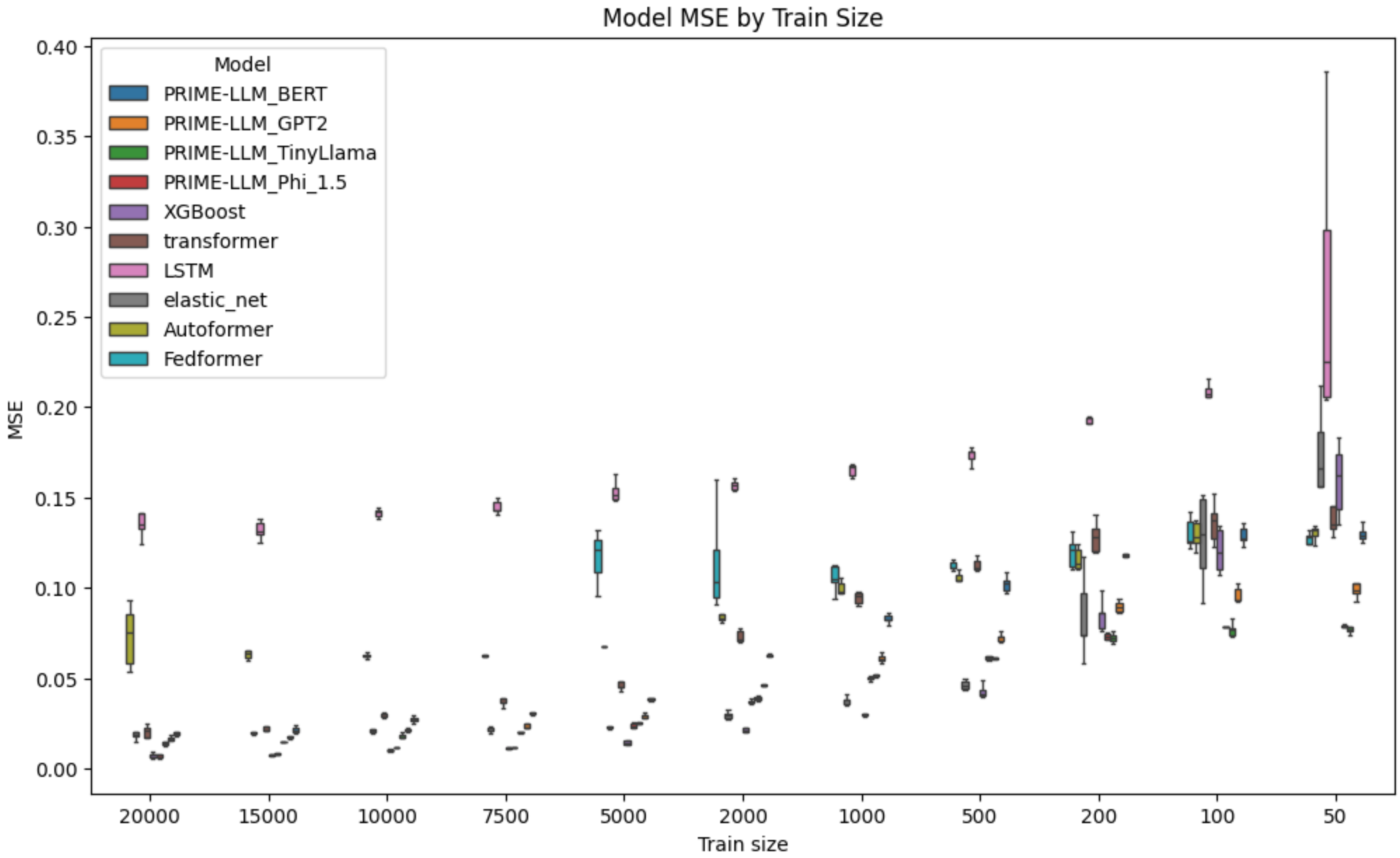


Supplementary Figure S1

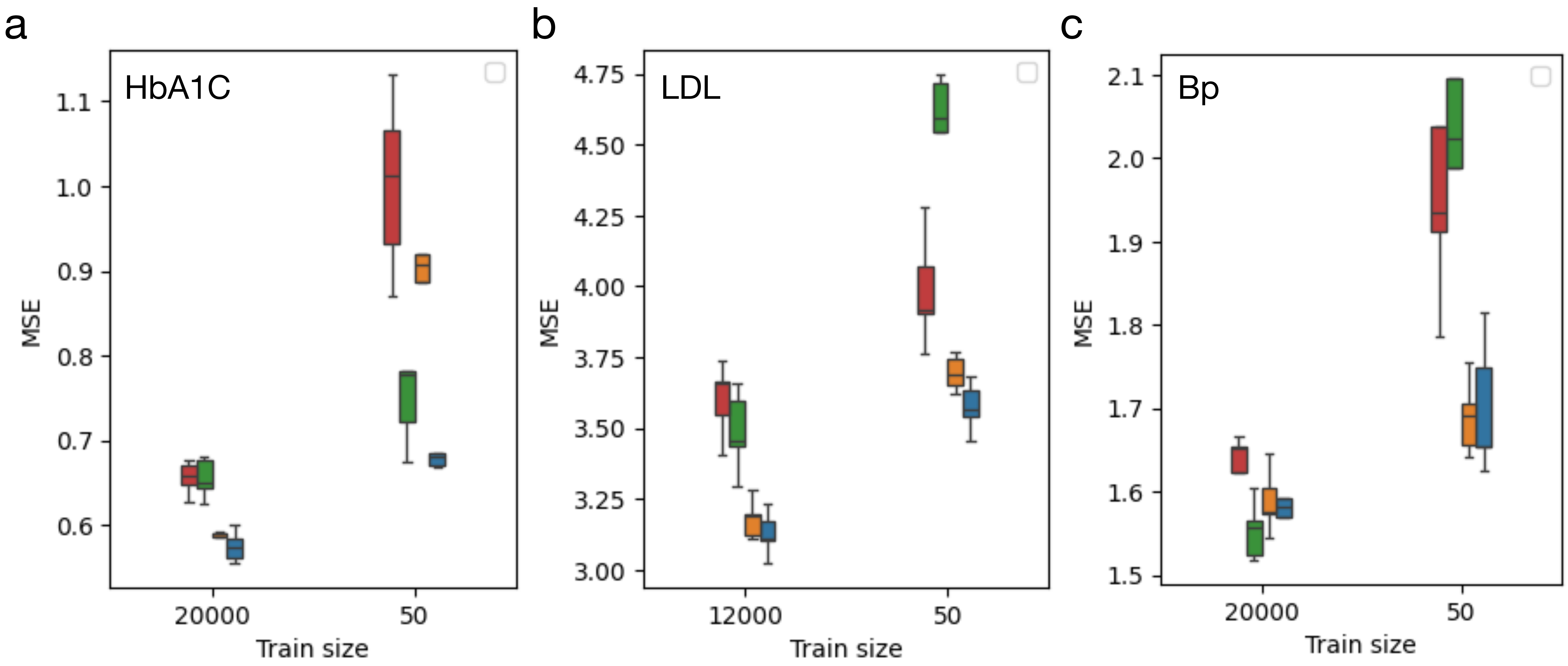


Supplementary Figure S2
